# Extensive Intra-host evolution and T cell escape by SARS-CoV-2 in a 2.5-year persistent infection of an immunocompromised host

**DOI:** 10.1101/2025.05.28.25328480

**Authors:** José Afonso Guerra-Assunção, Ruairi McErlean, Katie Townsend, Leonhard M Flaxl, Shengwei Jamie Tian, Leo Swadling, Judith Breuer, David M Lowe

## Abstract

Persistent SARS-CoV-2 infection in immunocompromised patients creates a unique environment for the accumulation of mutations linked to immune evasion. This is due to prolonged virus-host interaction, coupled with the effects of diminished selective pressures resulting from suboptimal adaptive immunity.

In this study, we investigated viral evolution and immune escape during a 2.5-year persistent SARS-CoV-2 infection in a patient with a history of multiple myeloma and rheumatoid arthritis on B cell depleting therapy. Sequencing of a SARS-CoV-2 virus isolate 899 days post-infection revealed a phylogenetic background compatible with the ancestral B.31 lineage. Nonetheless, the isolate displayed extensive intra-host evolution, evidenced by the accumulation of 56 non-synonymous mutations and one insertion across 20 viral proteins, as compared to the parental sequence. Apart from extensive private (infection-specific) mutations, the emergence of convergent mutations previously noted during other chronic infections and also in the later emerging variants of concern was observed.

SARS-CoV-2-specific humoral immunity was undetectable in the patient; however, despite long-term antigen exposure, cellular immunity remained high in magnitude, functional, and responsive to peptide recall, without driving T cell dysfunction/exhaustion. T cell memory was broadly targeting but dominated by CD8+ T cells recognising the spike protein. No non-synonymous mutations accumulated in B cell epitopes, as expected. However, 38/56 were present in 93 CD4 and CD8 T cell epitopes. Computational models predicted reduced MHC binding or immunogenicity for 72% (40/55) of the CD8 epitopes affected. Importantly, functional assays confirmed T cell escape at 50% (1/2) and 86% (6/7) of the CD8 and CD4 epitopes tested *in vitro*.

This study provides critical insights into viral adaptation and immune escape, demonstrating the ability of SARS-CoV-2 to explore a broad mutational landscape during long-term persistent infection, adapting to persistence and evolving within host to evade T cell recognition.

## Introduction

Acute SARS-CoV-2 infections typically resolves within two weeks, as the host immune system mounts effective innate and adaptive responses to clear the virus^1^. However, in immunocompromised patients, who fail to generate robust immune responses due to underlying primary immunodeficiency disorders, immunosuppressive medications, hematologic malignancies or other conditions, SARS-CoV-2 can persist for months or even years, leading to recurrent symptoms, progressive respiratory decline, and an increased risk of mortality^2^.

Prolonged infection allows ongoing viral replication under weak immune selection pressure in the host, during which time mutations can occur^3–7^. Many mutations are expected to be detrimental to the virus, but some can provide fitness advantages and may be selected for. This led to the hypothesis that prolonged infections in immunocompromised individuals may give rise to novel variants of concern, and that host-adapted and immune-evading variants can then be transmitted back to the population though prolonged shedding. Indeed, there is significant interest into whether the Omicron variant may have evolved during persistent infection in an immunocompromised host^8,9^.

Characterizing viral evolution and immunity during prolonged infections is crucial to understanding viral adaptation, immune evasion mechanisms (including T cell exhaustion and escape), the pathophysiology of COVID-19 and potentially mechanisms underlying Long COVID^3,10,11^.

In a previous study we characterised a set of clinical outcomes for immunocompromised patients receiving intravenous immunoglobulin (IVIG) treatment^12^. Here, we describe the extensive intra-host evolution of an isolate related to an ancestral B.31 lineage during a 2.5-year infection. We demonstrate that strong, high quality T cell memory is generated during persistent infection, dominated by spike-specific CD8 T cell responses. We do not identify mutations that would be expected to lead to resistance to monoclonal antibodies or antivirals, but we observe loss of recognition by host T cells of the emerging virus at several epitopes, demonstrating T cell escape.

## Results

### Clinical characterisation of a 2.5-year infection

A patient with a history of multiple myeloma and rheumatoid arthritis, treated with prednisolone (corticosteroid) and rituximab (anti-CD20 monoclonal antibody, mAb), suffered recurrent episodes of cough, breathlessness and fever from July-2020. Lung imaging demonstrated multifocal ground glass changes in a changing pattern (**Fig. 1, Supp Fig. 1**). SARS-CoV-2 PCR, including on bronchoscopy, was consistently negative. She was admitted to hospital in December 2022 due to acute respiratory deterioration and required intensive care admission. At this time, SARS-CoV-2 was identified on nasopharyngeal swab. The patient was receiving anti-CD20 medication and did not have detectable anti-spike antibody before IVIG treatment, despite multiple vaccinations and prolonged infection. She was treated with intravenous remdesivir (200mg on Day 1 then 100mg daily for a total of 10 days) and immunoglobulin (1g/kg over two days) with clinical recovery, detection of (passively acquired) spike antibody in serum and conversion of SARS-CoV-2 PCR tests to negative. She has since been maintained on replacement immunoglobulin therapy indefinitely but has ongoing breathlessness, wheezing and hypoxia, persisting lung imaging changes consistent with chronic airways disease and often superimposed infection (**Supp Fig. 1**), recurrent isolation of bacterial pathogens in sputum (*Haemophilus influenzae* and *Pseudomonas aeruginosa*) and frequent need for antibiotic therapy. Notably, prior to the SARS-CoV-2 infection she had no clinical or radiological lung disease. Blood tests reveal persistent lymphocytosis (up to 8 x 10^9^ cells/litre). She remains on prednisolone (maintenance dose 10-15mg daily) and has received further rituximab infusions 6-monthly.

**Figure 1:**
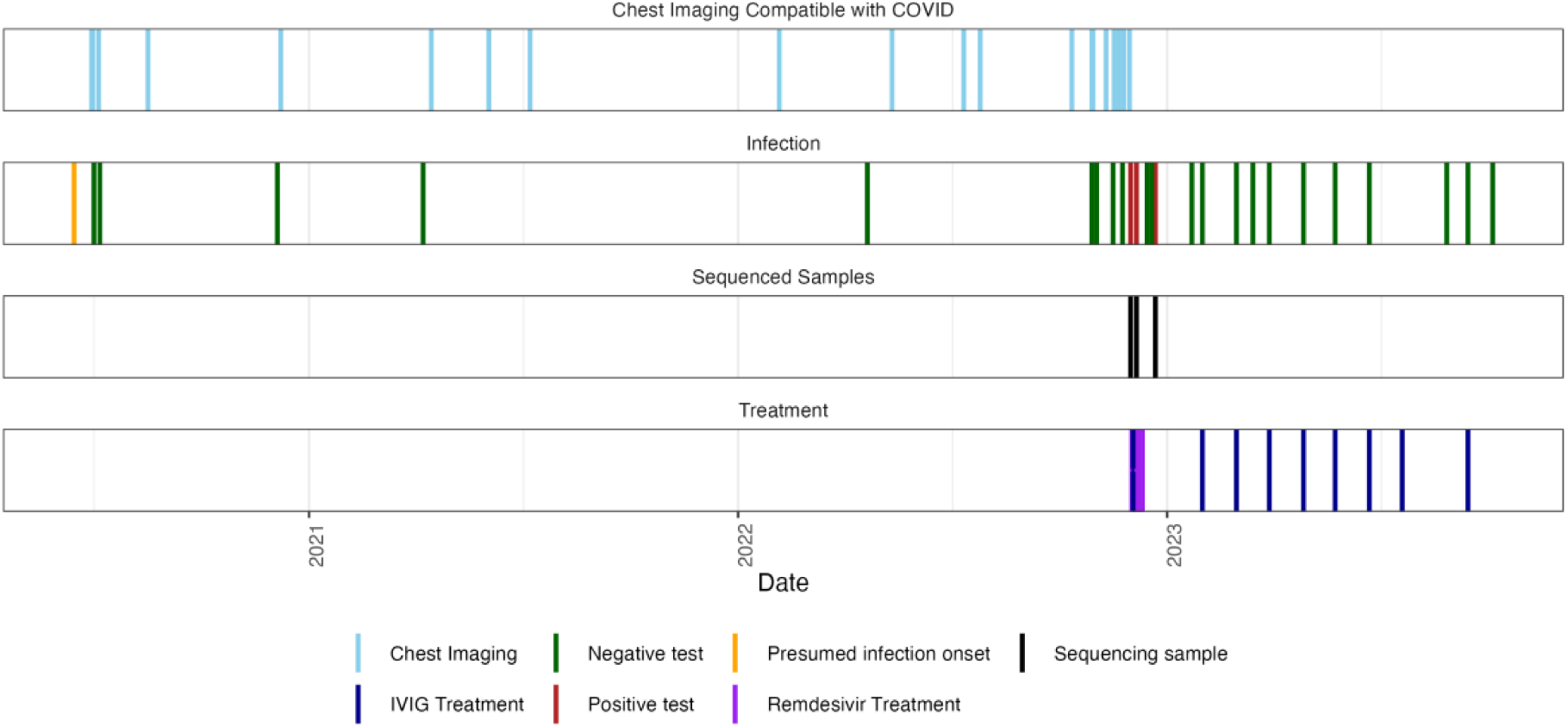
Timeline of clinically relevant events during persistent SARS-CoV-2 infection. Vertical lines indicate key clinical events over the 2.5-year infection period. Chest imaging: CT scans and chest X-rays demonstrating typical changes consistent with COVID-19 (**Supp** Fig. 1). Infection status: Negative (green) and positive (red) PCR tests on nasal swabs, sputum samples, and lateral flow tests. Treatment: Administration of intravenous immunoglobulin (IVIG, purple) and remdesivir (10-day course, magenta). Sequencing samples: Specimens collected for SARS-CoV-2 viral sequencing (**Supp Table.1**). The presumed infection onset is marked at the beginning of the timeline. Note persistent radiological changes despite intermittent negative PCR results throughout the infection period.

### Virus isolated during persistent SARS-CoV-2 infection accumulates convergent and infection specific mutations

Despite a clinical presentation indicative of SARS-CoV-2, virus was repeatedly not detected by Lateral Flow Test (LFT) or PCR on nasal swabs, sputum or even bronchoscopy, as has been described for other persistent infections^4^. Virus was isolated from this patient by nasal swab 899 days after suspected onset of SARS-CoV-2 infection (PCR cycle threshold 32). Sequencing quality control failed when viral loads were lower, with 1/3 samples from this patient passing QC filters for downstream analysis (**Supp Table.1**). UShER (Ultrafast Sample placement on Existing tRees^13^) was used to determine the rapid phylogenetic placement of the genomic sample within an existing reference 2 million SARS-CoV-2 phylogenetic tree using publicly available sequence data from GISAID (Global Initiative on Sharing All Influenza Data^14^). In agreement with the first onset of respiratory symptoms observed in the patient in mid 2020, the genome sequence was assigned as ‘Pango’ lineage B.31 (Emerging clade: B (19A), reassigned from B.2.5. Circulating in Africa, Asia, Oceania, North/South America and Europe [UK] from March 2020 to July 2020). UShER placement revealed the closest ancestral sequence as a virus isolate sampled from Wales in April 2020 (Wales/PHWC-28F23/2020|2020-04-02, EPI_ISL_445459). Only 283 virus genome records corresponding to the B.31 Pango lineage were available in the GISAID database (as of May 2025), predominantly sampled from the UK.

To determine the number of mutations accumulated in the genome, a map of single nucleotide polymorphisms (SNP) relative to the closest matching reference sequencing was created based on a pairwise comparison of the genetic identity between consensus genomes (**Supp Fig. 2**). In contrast to other shorter persistent infections we studied^12^, where the number of accumulated mutations observed relative to the closest related sequence were fewer than 15, this sample differed from its closest public B.31 relative by 79 mutations. Out of these, 56 were non-synonymous (**Fig. 2**). The virus had 5 mutations characteristic of ancestral viruses: ORF1ab:L3606F, ORF1ab:D54Y, ORF3a:G25IV, ORF8:S84L and N:D22G (**Fig. 2**).

**Figure 2:**
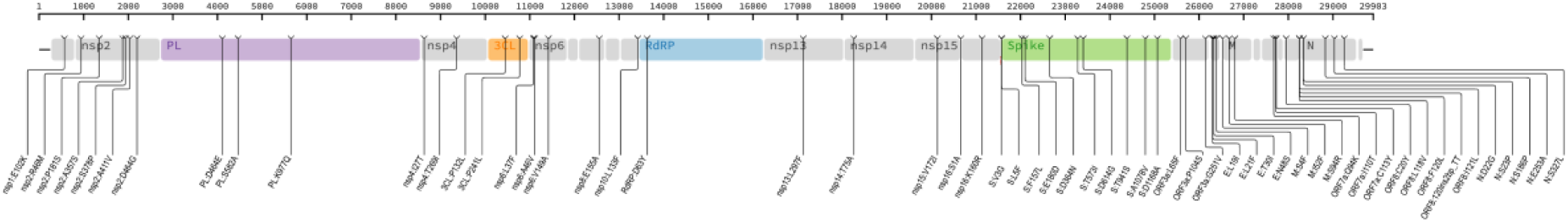
Distribution of non-synonymous mutations across the SARS-CoV-2 viral proteome at consensus level detected at day 899 of infection: Amino acid position shown above. Mutations listed below as affected protein, ancestral amino acid, position in protein, and variant amino acid. Mutations in spike highlighted in bold. 3CL, 3C-like protease, E, envelope; M, membrane protein; N, Nucleoprotein; NSP5; NSP, non-structural protein; ORF, open-reading frame; PL, papain-like protease NSP3; RdRP, RNA-dependent RNA-polymerase.

Despite intra-host evolution, the treatment regimen administered (remdesivir plus IVIG) was sufficient to finally control the infection, as assessed by sustained viral clearance post treatment (**Fig. 1**). This patient was subsequently re-infected with a XEC.2 strain of SARS-CoV-2 in October 2024 (ENA Biosample: SAMEA117560877). The virus isolated from this subsequent infection did not share any mutations or regions of similarity suggestive of recombination or further persistence of the virus isolated during primary infection.

### 2.5-year infection leads to extensive mutations across the viral genome

We first investigated whether the genetic variation seen within the sequence taken at day 899 could have been the result of viral recombination during the persistent infection. At the time of sequencing (Dec 2022) the dominant variants circulating in the UK and Europe were BA.5, BQ.1 and XBB. We found no evidence of contiguous mutation stretches or lineage defining mutations shared between the patient sample and globally circulating viruses (**Fig. 2, Supp Fig. 2**). This finding militates against viral recombination with newer reinfecting variants as the origin of the polymorphisms observed, instead strongly suggesting that viral genetic variation within this individual evolved during chronic infection.

Next, we considered whether the mutations arising in our patient were common to persistent infections, indicative of convergent evolution. Two studies have compiled data from several sources on the mutations acquired during persistent infections that may indicate evasion of adaptive immunity or adaptation of the virus in the specific setting of chronic infection (where selection pressures differ to acute resolving infections)^10,15^. Of the 21 mutations that have been observed in multiple persistent infections (168 genomes, as compiled in^15^) two emerged in the patient described here: E:T30I and NSP3:K977K (also observed as a lineage defining mutation for the P.1 gamma VOC). Wilkinson *et al* describe the E:T30I mutation as a “sensitive marker” of persistent infection due to its repeated appearance in different studies and no association with a particular source, geographical region, or SARS-CoV-2 lineage^15^. Mutations within the spike protein, V3G and A1078V seen in our patient have also been reported previously in persistent infection^10,11^. The most comprehensive study of persistent infection using repeated PCR positivity also identified ORF8 mutation I121L as being recurrently identified in prolonged infections^16^. Overall, a subset of the amino acid changes arising during the persistent infection have been previously described, suggesting convergent evolution that might confer a fitness advantage in this setting.

Some of the non-synonymous mutations acquired during this persistent infection are rarely observed elsewhere globally (16 mutations observed in <1000 sequences sampled globally since the beginning of the pandemic to May 2025), however, several are characteristic of VOCs or sub-lineages that subsequently emerged, such as: S:D614G, NSP3:K977Q, NSP6:V149A, S:F157L, S:T573I, ORF3a:P104S, NP:S327L. In particular Spike D614G mutation has been widely studied, marks the B.1-like genotypes^17^, and was present in almost all sequences post-May2020, but was lacking in B.31. This suggests that features of emerging variants in the general population may be predicted from studies of intra-host evolution in chronically infected individuals^18^.

The spike protein is the main target of humoral immunity, in particular for neutralising antibodies, but is also immunodominant within T cell responses seen after infection^19^. This protein also plays a key role in viral entry to the host cell, and it is therefore the main viral protein undergoing positive selection. Analysis of the frequencies of the spike mutations acquired in our patient within the GISAID database revealed that most of the nine fixed NS spike mutations were not widely detected in general circulation (mutations E180D, D364N, A1078V and D1168A were observed in approximately 62, 3, 125 and 7 sequences per million, respectively), suggesting they may reduce viral fitness^20^. The spike mutations V3G, S:F157L, T573I, and D614G, conversely have circulated more widely, however, in no other publicly available sequence have these 4 mutations been observed together.

In this seronegative patient, mutations in structural proteins were not associated with antibody escape, and mutations were not enriched in the receptor binding domain, the main target of neutralising antibodies. Mutations did not appear to be randomly distributed across viral proteins, however, as they were less common than expected in the non-structural proteins (NSPs) of ORF1ab, which accounts for ∼71% of the viral genome but contained just 45% of the mutations. This observation is in line with the notion that non-synonymous mutations are less tolerated on non-structural proteins, as these are likely purged through purifying selection, due to functional and structural constraints in proteins that play a vital role in viral replication^21,22^

As described above, no protease or polymerase inhibitors or SARS-CoV-2-specific mAb were administered prior to the virus being isolated at day 899 of infection. As expected, none of the 9 mutations in spike observed during persistent infection have been described as mAb resistance mutations (Stanford CoV-RDB^23^). Only mutation D364N falls within the RBD but it does not directly disrupt known RBD antibody binding sites, and as such is not predicted to cause antibody escape^24^. The mutations that fall in NSP5 (3CLpro) and NSP12 (RdRp) have not been linked to resistance to 3CLpro or polymerase inhibitors^23^.

Overall, these data suggest that a B.3 virus (early Wuhan-hu-1-like isolate) has undergone extensive evolution during an infection that has lasted over 2.5-years, leading to recurrent emergence of mutations that are common in persistent infections and in subsequently emerging VOCs, but also a large set of unique (private) mutations.

### Robust and functional T cell memory is established despite persistent infection

Chronic viral infections and associated persistent antigen exposure can drive T cells to become dysfunctional and ‘exhausted’^25^. SARS-CoV-2 is not adapted to persist in immunocompetent individuals, but whether persistence in immunocompromised patients leads to virus-specific T cell exhaustion has not been well studied.

We next assessed what T cell immunity was generated by this long-term persistent infection without seroconversion. The memory T cell response targeted all SARS-CoV-2 proteins tested, but was strongest to the spike protein (**Fig. 3a**). When quantifying the SARS-CoV-2-specific T cell response in the blood, the total magnitude was larger than that detected in pre-pandemic samples, in exposed-seronegative healthcare workers^21^, and larger than all but one response seen 4 months after infection with the emergent Wuhan-hu-1 sequence (**Fig. 3b**, n=71), despite it being measured here ∼ 1 year after resolution of infection and at the time of corticosteroid use. This high magnitude T cell response was dominated by structural protein-specific T cells, as has been described for memory responses after infection with Wuhan-hu-1 SARS-CoV-2^26,27^ (**Fig. 3c**); however, a T cell response to the non-structural proteins that make up the core of the viral replication-transcription complex (RTC) was also seen. RTC-specific T cells have been associated with early control of viral replication, being enriched before and after abortive SARS-CoV-2 infection both in natural history studies and human challenge^21,28^. The ratio of RTC- to structural protein-specific T cells was lower during persistent infection than seen after an acute-resolving Wuhan hu-1 infection, suggesting prolonged infection biases the T cell response towards spike and structural proteins (**Fig. 3d**). This patient had a high frequency of total CD8+ T cells (66.0% of CD3+, versus CD4+ 29.4% of CD3+) and a low frequency of naïve T cells (**Supp Fig. 3a-c**). The T cell response to control antigens from Influenza (Flu), EBV and CMV was also relatively high in magnitude, in the 97th percentile of all healthy donors tested, which may be due to a higher frequency of T cells in the blood relative to other PBMC subsets for this immunocompromised patient (1678 SFU/10^6^ PBMC; **Fig. 3e; Supp Fig. 3a**).

**Figure 3:**
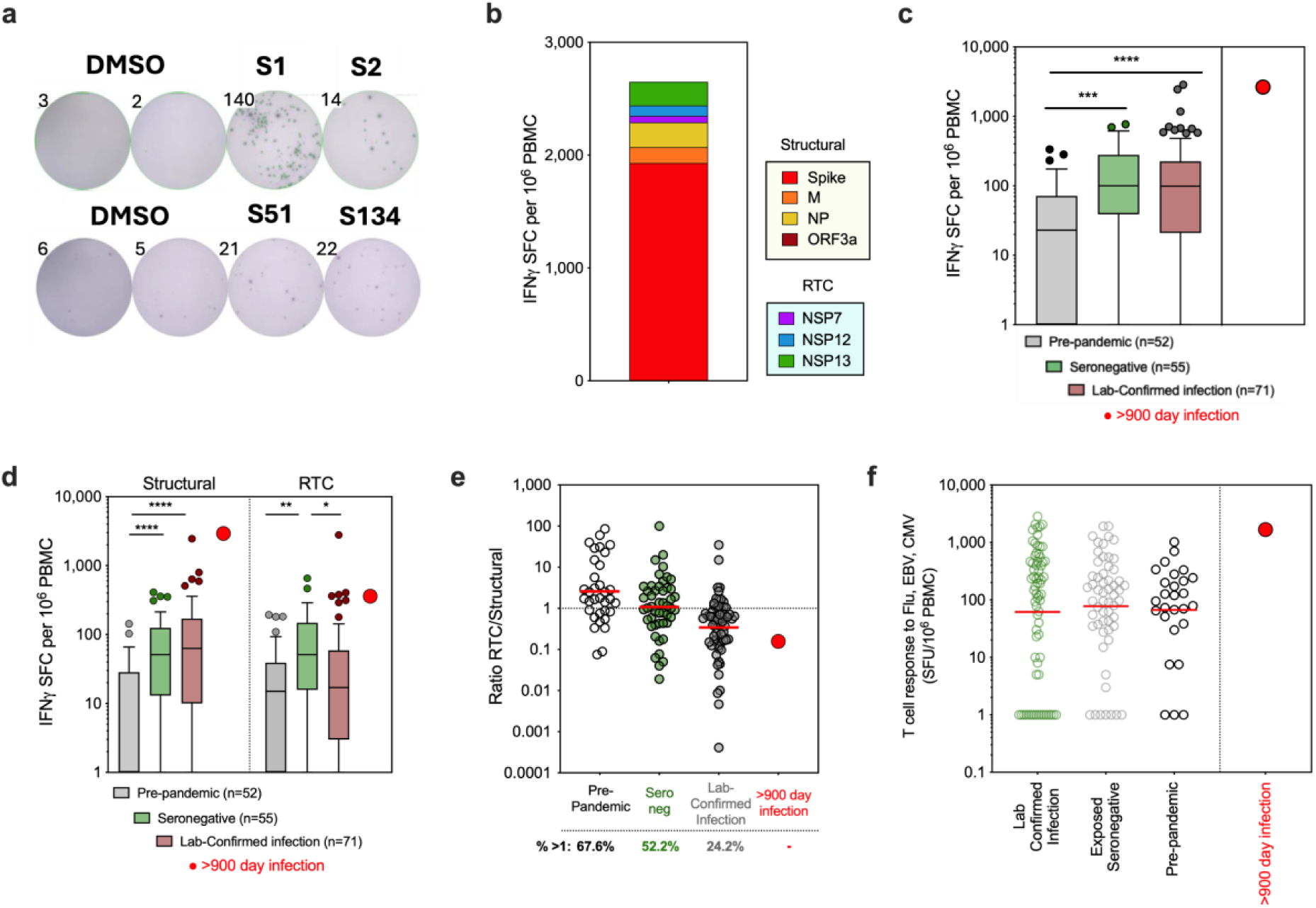
Magnitude and specificity of T cell memory responses generated by 920-day infection. **a)** *Ex vivo* IFNγ-ELISpot well images. DMSO control unstimulated wells and wells stimulated with overlapping peptides covering S1 and S2 region of Spike, or single peptides S51 and S134 shown. **b)** Total magnitude of SARS-CoV-2-specific memory T cell response to structural proteins Spike, membrane (M), nucleoprotein (NP) and open-reading frame 3a (ORF3a) and replication-transcription complex (RTC) proteins (NSP7, NSP12 polymerase, NSP13 helicase) coloured by protein targeted and measured after persistent infection (>900 days). **c-d)** Total magnitude of SARS-CoV-2-specific T cell response **c)** or T cell response to Structural and RTC proteins **d)** in pre-pandemic samples (pre-August 2019), in exposed healthcare workers (HCW) who remained seronegative, including abortive infections, and in HCW with laboratory confirmed SARS-CoV-2 infections (samples 4 months post-exposure/infection in Jun-July 2020) for comparison to T cell response in persistently infected patient. **e)** Ratio of the magnitude of the T cell response to RTC/Structural T cells. Percentage of cohort with a response above 1 (stronger response to RTC than structural proteins) shown below. **f)** Magnitude of T cell response to a pool of epitopes from Flu, EBV and CMV. **(b-e)** subset of data previously published in Swadling *et al*^21^. **a-f**) IFNγ-ELISpot. **c)** Box and Whisker, Tukey. **c-f)** statistical analysis was performed using Kruskal–Wallis tests with Dunn’s correction.

We next used surface marker and intracellular cytokine staining (ICS) to determine whether these SARS-CoV-2-specific T cells were CD4+ or CD8+, and to confirm their functional capacity. ICS confirmed the immunodominance of spike relative to non-structural proteins (**Fig. 4a**; Gating strategy **Supp Fig. 3a)**. Spike-specific T cells were predominantly CD8+ (∼2.73% of CD8 and ∼0.36% of CD4+ T cells IFNγ+) and targeted the S1 region. CD4+ T cells to both S1 and S2 regions were also detected. T cell responses to RTC were lower in magnitude but more balanced between CD4 (∼0.06% IFNγ+) and CD8 (∼0.05% IFNγ+)(**Fig. 4b**). SARS-CoV-2-specific CD4+ and CD8+ memory T cells were highly functional, producing the antiviral and immunomodulatory cytokines IFNγ, TNF and IL-2, expressing CD154 and degranulating (CD107⍺+) upon peptide stimulation *in vitro* (**Fig. 4c**). SARS-CoV-2-specific T cells were almost exclusively effector memory phenotype (Tem; CD45RA-CCR7-; **Supp Fig. 3d**).

**Figure 4:**
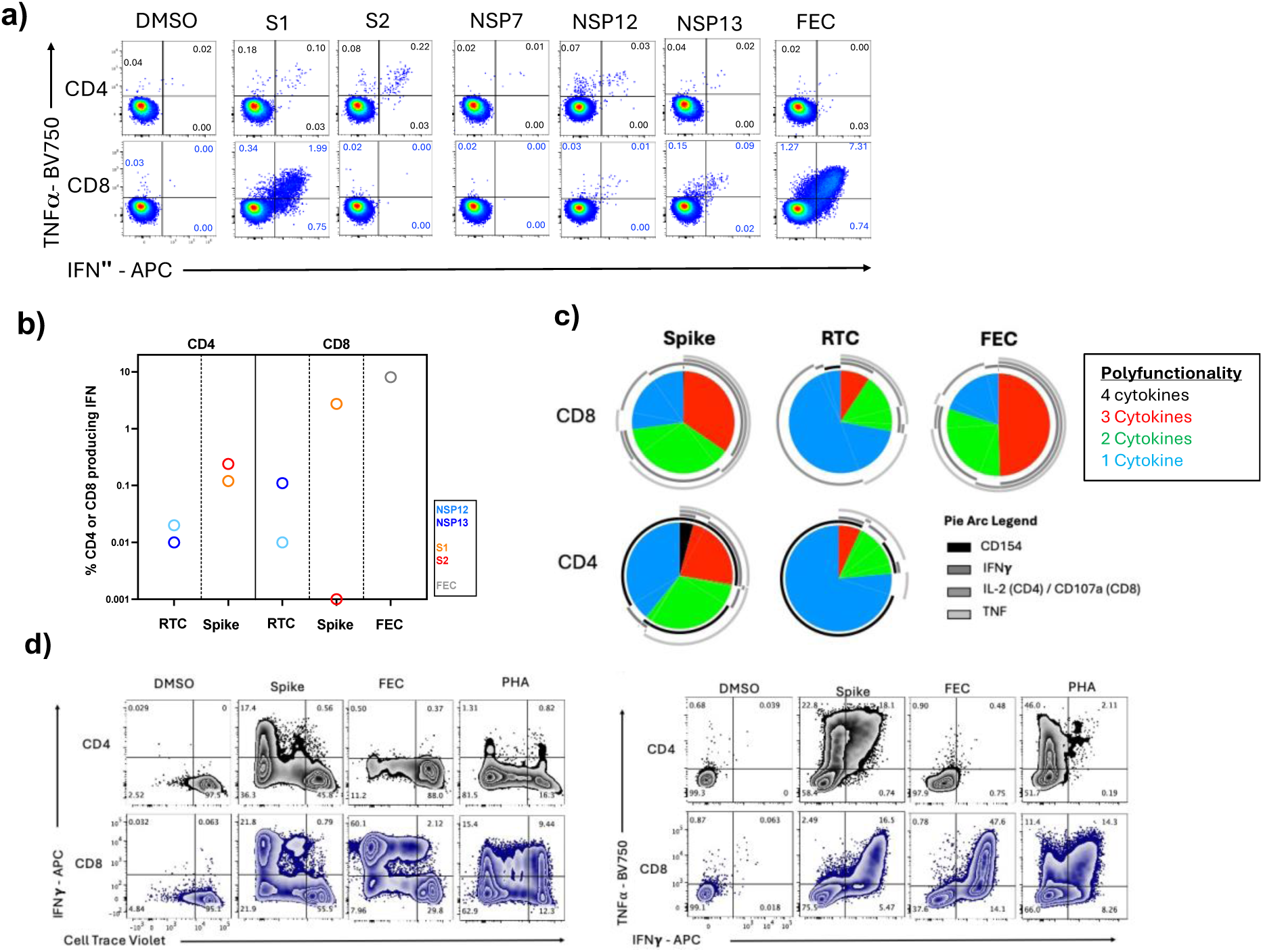
*Ex vivo* functionality and proliferative potential of T cell memory generated by a 920 day persistent infection. a) The IFNγ and/or TNF⍺ producing CD4+ and CD8+ T cell memory response to SARS-CoV-2 proteins after persistent infection. Percentage of CD4+ or CD8+ shown. Spike divided into S1 and S2 peptide pools. DMSO, unstimulated control. FEC, response to Flu, EBV and CMV epitope pool. b) Summed CD4+ or CD8+ IFNγ+ response to spike (S1, S2) or the replication-transcription proteins (RTC; NSP7/12/13). c) The proportion of SARS-CoV-2-specific CD4+ or CD8+ T cells to spike or RTC that co-produce a number of cytokines/effector molecules are shown as pie charts. Pie arcs show the proportion producing each single cytokine. d) Proliferation and expansion of SARS-CoV-2-specific memory CD4+ and CD8+ T cells post-persistent infection when *in vitro* stimulated with Spike, Flu EBV and CMV epitopes, or PHA, as shown by CTV dilution and IFNγ production after 8-day stimulation. Percentage of CD4+ or CD8+ shown for each quadrant. PHA, Phytohemagglutinin.

We next assessed T cell proliferative capacity. Both CD4+ and CD8+ SARS-CoV-2-specific T cells generated by persistent infection showed significant expansion *in vitro* in response to spike peptide pool stimulation (∼55-fold for CD4+, ∼8.3-fold for CD8+ for the IFNγ+TNF+ population comparing *ex vivo* cells to *in vitro* expanded cells), similar to the CD8+ T cell response to influenza, EBV and CMV (∼6.5-fold for CD8 IFNγ+TNF+, **Fig. 4d**).

Overall, this indicates that there was no functional exhaustion related to persistent antigen exposure in this long-term infection; rather, persistent infection leads to high magnitude, broad and functional T cell memory.

### Mutations acquired during persistent infection are predicted to impact the presentation and immunogenicity of T cell epitopes

Two major mechanisms by which variable viruses escape T cell recognition are: **1)** the introduction of mutations that reduce or abrogate peptide binding to MHC, reducing presentation of viral peptides to T cells (in particular mutations at 1,2 and c-terminal residues); and **2)** by introducing mutations that reduce TCR recognition of peptide-MHC complexes. To determine if the mutations acquired here would impact T cell recognition of virally infected cells we first determined if the 56 mutations fall within known T cell epitopes. A third of the mutations (18/56) had no impact on known epitopes, however, 38 mutations fell within 38 CD4 (**Supp Table.2**) and 55 CD8 epitopes (**Supp Table.3**).

We used machine learning tools to predict the impact of mutations on peptide presentation and immunogenicity. Mutations arising during this long-term persistent infection are predicted to negatively impact peptide binding and presentation to CD8+ T cells for 35% (23/66) of the peptide-MHC interactions, with 60% being unaffected and 5% predicted to improve peptide binding (**Supp Table.3; Supp Fig. 4**). Prediction of peptide binding to MHC class II is less accurate with current models. Only 13 of the validated SARS-CoV-2 epitopes were predicted to be binders for MHC class II **(Supp Table.2)**; of these, 6 were unaffected by the acquired mutations, 1 was negatively impacted and 5 had improved peptide binding. The mutations acquired during this persistent infection negatively impacted immunogenicity scores for 40% (22/55) of the CD8 T cell epitopes (**Supp Table.4**).

Overall, the mutations arising during this persistent infection are expected to negatively impact T cell recognition by reducing presentation and/or immunogenicity for 72% (40/55) of epitopes in which mutations fall.

### Mutations acquired during persistent infection led to T cell escape

To determine if the mutations acquired during the persistent infection led to functional loss of recognition and T cell escape, we took a subset of immunodominant epitopes in which mutations arose and measured whether the patient had a detectable T cell response to these. As expected, T cell responses to individual epitopes were low magnitude *ex vivo* (**Fig. 5a),** however, functional T cells could be readily expanded *in vitro* for a subset of these epitopes (**Fig. 5b).** Next, we tested whether the patients T cells could recognise the mutant versions of each epitope that had arisen during persistent infection. We expanded T cells with a pool of 12 peptides of ancestral or variant sequence, showing that the magnitude of the response to the ancestral infecting virus sequence was higher than the response to the variant peptide pool corresponding to the virus sequence at day 899 of persistent infection (**Fig. 5c)**.

**Figure 5:**
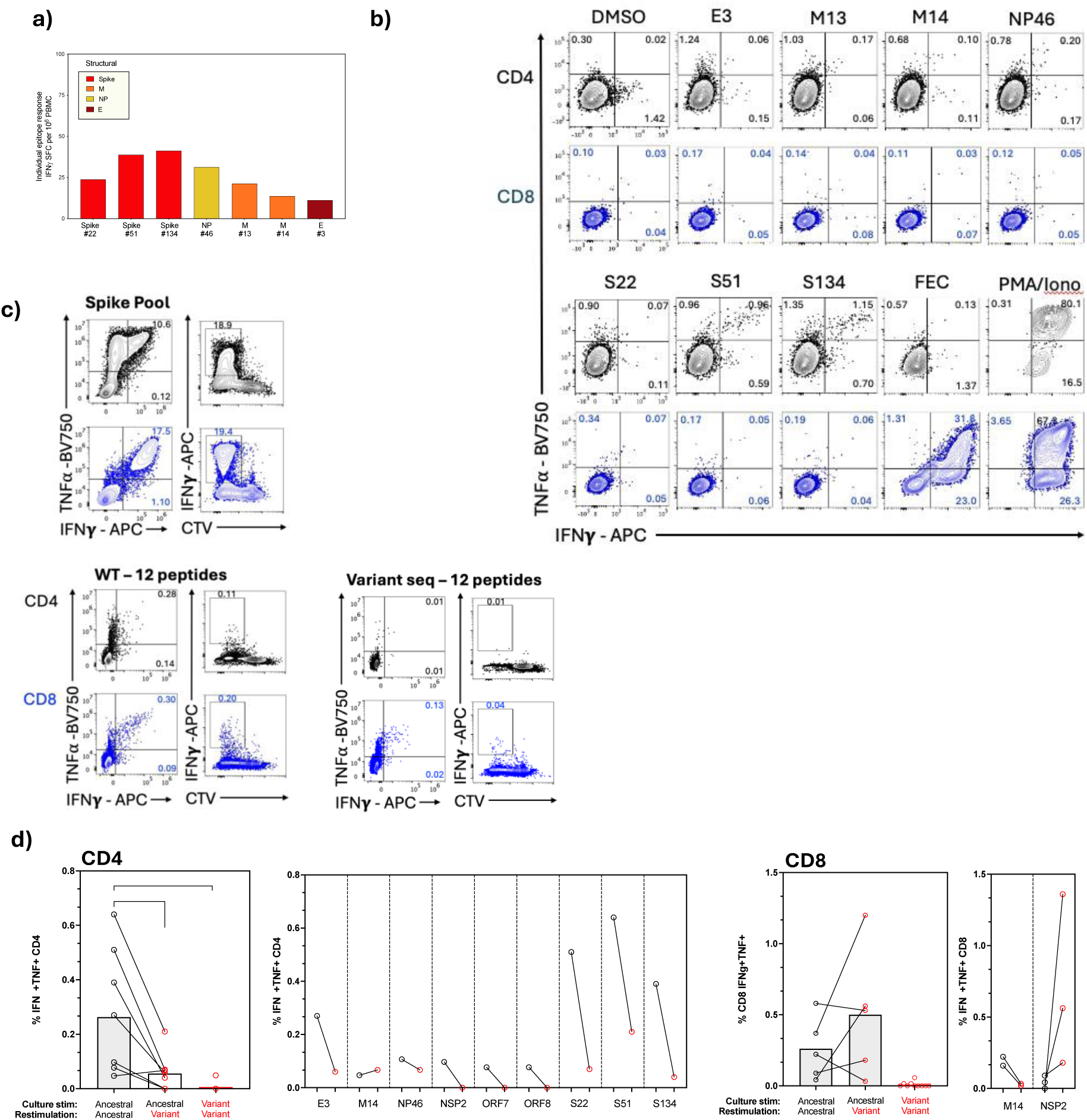
Failure of host T cells to recognize emerging virus. a) *Ex vivo* magnitude of the T cell response to individual ancestral sequence peptides in which mutation arose over persistent infection by IFNγ-ELISpot. b) Magnitude of the CD4+ and CD8+ T cell responses after 10-day *in vitro* peptide expansion with individual ancestral sequence peptides. Percentage of CD4+ and CD8+ producing IFNγ, TNF, or both are shown. c) Magnitude of IFNγ+, IFNγ+TNF+ and CTV^lo^IFNγ+ CD4+ and CD8+ T cells after 8-day expansion with a pool of 12 peptides corresponding to ancestral sequence epitopes or variant sequence epitopes containing mutations in the virus isolated at day 899 of infection. d) Magnitude of the IFNγ+TNF+ CD4+ or CD8+ T cell response after 10-day *in vitro* peptide stimulation with individual epitopes using ancestral sequence or variant sequence peptides for culture and restimulation on day 9. Summary data showing all T cell responses that were detectable (left) or shown for individual peptides (right) for CD4+ then CD8+ T cells. Duplicate and triplicate stimulations are shown for CD8+ T cells for M14 and NSP2 respectively. d) statistical analysis was performed using Kruskal–Wallis tests with Dunn’s correction.

When considering individual epitope responses, for 6/7 of the CD4 and 1/2 CD8 epitopes T cell recognition was almost completely abrogated against the variant peptide (**Fig. 5d**). The mutant version of M13 was, however, equally well recognised by CD4+ T cells as the ancestral sequence. A mutation in NSP2 (A411V; TSDLATNNLVVM**V**Y) lead to better CD8+ T cell recognition *in vitro* (**Fig. 5d**). Attempted T cell expansion *in vitro* with the mutant version of these epitopes, however, did not lead to T cell expansion except for CD4+ T cells recognising M13 and a very low magnitude CD8+ T cells response to NSP2 variant (0.01% variant expansion vs. 0.7% of CD8+ T cells on average for expansion with ancestral peptide; **Fig. 5d)**, confirming the lack of recognition of most mutant variants and demonstrating that mutations did not introduce *de novo* epitopes. Therefore, mutations arising during a 2.5-year infection led to loss of epitope recognition and T cell escape.

## Discussion

Here we have identified the longest SARS-CoV-2 infection described to date, persisting over 2.5-years based on clinical symptoms, lung pathology, and viral sequence characterisation despite the virus being undetectable in respiratory samples for the majority of the infection.

Persistent SARS-CoV-2 infections that last so long are rare and tend to occur in people with significant immunocompromise, with the most comprehensive study to date identifying just 10 of ∼94,000 infections lasting more than 100 days and none longer than 193 days when relying on PCR positivity to determine duration of infection^16^. It is important to note that it is still rare for patients on anti-CD20 therapy to develop persistent infection, with innate immunity and T cells sufficient to mediate control in most cases^29,30^, and so additional factors likely contribute to the inability of this patient to naturally resolve SARS-CoV-2 infection.

It is difficult to identify and study persistent infections that do not result in detectable virus in bronchoalveolar lavage or nasal samples, however, post-mortem studies have identified persistence of virus in bronchial glands and chondrocytes, without detection in respiratory epithelium, in individuals who died due to pneumonia^31^. Viral rebound is common in persistent infections^16^, and could be the result of acquired adaptation and immune evasion. This may also explain why detectable virus was only seen at day 899 in our chronic infection despite regular testing and constant clinical indicators of infection. This highlights the importance of using a mixture of molecular and clinical tools to identify and study viral persistence, especially given the risk of irreversible lung damage as seen in our patient.

The rate of mutations observed during persistent infections has been estimated to be ∼2x higher than that seen when tracking at the population level^4,11,32^. Transmission is characterised by a narrow bottleneck^33^ and a period of low mutation rate has been observed in ‘short’ persistent infections in immunocompetent patients (56-100 days^16^), suggestive of weak selective pressures. In contrast, persistent infections >100 days, almost invariably in immunocompromised patients, releases the virus from the constraints of transmission bottlenecks and give the opportunity to adapt and acquire immune escape mutations. As the longest SARS-CoV-2 infection described to date, an extensive mutational profile was observed, with 79 mutations, resulting in 56 non-synonymous and one insertion affecting almost all viral proteins. These mutations were indicative of an ancestral strain that had acquired an extensive array of mutations. The virus mapped to the B.31 lineage, which had not been identified since July 2020, and importantly it showed no signs of recombination or characteristic mutations of the variants circulating at the time of isolation. It carried several key mutations of ancestral stains (ORF1ab:L3606F, ORF1ab:D54Y, ORF3a:G25IV, ORF8:S84L and N:D22G), a set of convergent mutations that had been previously seen only in persistent infections (E:T30I and NSP3:K977K ORF8 mutation I121L^15,16^), as well as a set of unique or private mutations.

Persistent infection can offer a window into how SARS-CoV-2 may evolve at an inter-host or population level, with this viral isolate acquiring several mutations that became fixed in later variants (in particular S:D614G and NSP3:K977K). The now almost universal D614G mutation in spike enhances viral transmission by increasing S protein stability and binding affinity^17,34^. The absence of D614G in the closest sequence to this sample suggests that this infection occurred when D614G was not yet ubiquitous, although it is impossible to confirm if it appeared independently in this patient, as the data suggest. As immunocompromised patients have been suggested as the origin of VOCs^16,35^ this underpins the importance of monitoring their infections for viral clearance^36^.

An important consideration for viral evolution in antibody-deficient patients is that the lack of humoral immune pressure reduces selective forces shaping the virus. Without an effective antibody response, mutations enabling antibody escape do not provide a replicative advantage. However, some immunocompromised patients retain marginal B cell function sufficient to exert weak antibody pressure^37^ or might receive inadequate titres of antibody via therapeutic products. Here we saw no mutations that have previously been associated with antibody evasion and no enrichment for mutations in spike and its receptor binding domain, in line with a complete lack of SARS-CoV-2-specific humoral immunity prior to virus isolation. Our analysis revealed that most fixed spike mutations emerging in this chronically infected patient are likely to reduce viral fitness as they are rare in global sequence databases. Structural studies indicate two of the mutations arising,T573I and A1078V, may destabilize the structure of the spike protein^10,38^.

Long-term persistent infection did, however, induce a high magnitude, broadly targeting, proliferative, and functional memory T cell response. SARS-CoV-2-specific T cell memory persisted to a year after viral control at levels that are higher than those seen in immunocompetent patients 4 months after wuhan hu-1 infection^21,27^. Interestingly, rapid production of multiple antiviral cytokines and good proliferative capacity of the generated T cell memory shows that there were no signs of functional T cell exhaustion despite persistent antigen exposure for 2.5-years. Viruses that have evolved to allow chronic infection in humans can alter the immune landscape during infection and drive exhaustion via multiple mechanisms^25,39–41^, however, antigen exposure alone does not always drive exhaustion, as is observed in autoimmunity and latent infections such as EBV and CMV, which could explain the lack of exhaustion here.

The expansion of a large, spike-dominated T cell response is characteristic of more severe SARS-CoV-2 infections; it has been suggested that this skewing towards a spike-specific immune response is the result of higher and longer antigen exposure^42,43^. Even in mild infections, those with the lowest Ct value by PCR (i.e. highest viral load) tend to generate a more spike-dominated T cell response^21^. T cell responses recognising the non-structural proteins of the core replication-transcription complex (RTC) have been associated with early viral control and protection from detectable infection^21^. A detectable response to these proteins was observed in our patient despite failure to clear the virus, however, the ratio of RTC:Spike T cells was lower than that observed in acute-resolving infection, showing that they remain subdominant. The RTC proteins in ORF1ab are highly conserved, likely due to functional constraint and their importance in the viral life cycle^21,22^. Despite ORF1ab constituting more than two-thirds of the overall SARS-CoV-2 genome by length, only 45% of the mutations arising in our patient were within ORF1ab, suggesting that they acquired mutation more slowly than other ORFs. The exact impact of mutations in ORF1ab are hard to determine due to a lack of tractable models to study them *in vitro*, unlike for receptor binding and antibody evasion by changes in spike.

At the population level, the diversity of HLA molecules ensures that each individual recognises a different set of epitopes across a virus, which spreads the selection pressure and makes it more difficult for the virus to accrue T cell escape mutations. This also makes it difficult to see positive selection at T cell epitopes. For influenza, positive selection can be observed by comparing evolution at HLA presented epitopes isolated in parallel in humans and pigs (where there is no presentation and selection pressure)^44^. Long-term T cell escape occurs over decades^45^ and in mouse models escape mutations revert back to wild type in a host lacking the HLA to present the escaped epitope^46^. Intra-host escape from T cells is, however, well described for viruses that commonly cause chronic infections, such as HIV, and hepatitis B and C^40,47,48^. The selection pressure from T cell immunity persists throughout the infection at the specific epitopes presented by the patient’s HLA, which can drive the fixation of mutations in known T cell epitopes. Here, 72% of the CD8 epitopes in which mutations fell were expected to be negatively impacted by *in silico* prediction of peptide MHC binding and immunogenicity. Most importantly, we have shown that mutations arising within both CD4 and CD8 epitopes for which T cell responses are seen, result in lack of recognition when T cells are stimulated with the emerging variant sequences, demonstrating functional T cell escape. The magnitude of the T cell response to these 12 epitopes is, however, low and just a fraction of the total T cell response in this patient. Many epitopes were retained despite persistent infection and the resulting mutant virus had lost a small subset of the total epitopes seen across the SARS-COV-2 genome. Individual peptide responses were more readily detectable for CD4+ responses and only 1 of the two CD8 epitopes showed escape, versus 6/7 CD4+ responses. Lineage defining mutations for SARS-CoV-2 variants impact just a small number of epitopes and lead to a small but consistent drop in T cell recognition^49^. We have previously shown that despite most lineage defining mutations falling in T cell epitopes, this is not occurring more than by chance, suggesting that selection pressure by T cells is unlikely the main driver of evolution of these mutations^49^. As with the mutations arising in emerging variants^50,51^, the mutations arising during this persistent infection affected just a subset of epitopes, leading to a small but detectable loss of T cell recognition. This suggests that persistent infection offers a window into the future of potential escape mechanisms seen in variants of concern.

It is not possible to determine whether the T cell response in this patient was responsible for keeping viral replication below that which is detectable for the majority of the infection, and/or whether T cell-driven inflammation is responsible for the ongoing lung pathology observed. In post-mortem analysis of persistent infections, lung pathology at the time of death was similar to that seen at the time of PCR positivity in acute SARS-CoV-2 infection and NP and spike antigens were readily detectable^31^. Additionally, persistent infections in immuno-competent individuals have been linked to higher rates of Long-COVID^16^; a better understanding of immune-surveillance and mechanisms of immune escape in persistent infections may shed light on the pathophysiology of Long COVID and highlight therapeutic targets, as well as informing clinical management of immunocompromised individuals.

This study may have limited generalizability due to the fact that it is a single case report. The absence of detectable virus in samples collected throughout the 2.5-year infection period prevents a detailed analysis of the evolutionary trajectory. Although the GISAID database represents the most extensive collection of viral sequences over time and geographical regions, a major caveat when comparing the frequency of mutations is that there is inherent sampling bias due to uneven sequencing efforts. Additionally, blood samples for immunological assessment were taken more than one year after resolution of the infection, which may have affected the T cell responses we detected. We were unable to determine the patient’s HLA type, which limited our ability to precisely predict epitope binding and T cell recognition. Our investigation covered only a subset of T cell epitopes affected by viral mutations, providing an incomplete picture of the total immune escape landscape.

Overall, the viral evolutionary pathways in immunocompromised patients highlight the potential for SARS-CoV-2 to explore a wider mutational landscape in the absence of full immune constraint. However, without significant humoral immune escape mutations, it is likely that antibody-based treatment (i.e. IVIG, convalescent plasma or effective monoclonal antibodies) would be effective in these patients in combination with antivirals. This work highlights the importance of close monitoring immunocompromised patients, where infections may not always be detectable by nasal swab and RT-PCR, but where therapeutic intervention may be required to limit progression of lung pathology. While this study describes a single infection, the findings suggest that T cell-mediated selective pressure may be selecting epitope variants that confer immune evasion. Larger studies will be required to fully elucidate the prevalence and phenotypic consequences of this phenomenon. Nonetheless, our work highlights the potential importance of monitoring T cell reactivity against mutated epitopes over time, as changes in epitope recognition may serve as an early indicator of viral immune escape. Integrating longitudinal genomic and immunologic data from larger cohorts promises to shed further light on the interplay between the human immune system and evolving viral genomes.

## Data Availability

All data analysed during this study are included in this published article and its Supplementary Information.
Viral sequences are available at: European Nucleotide Archive (ebi.ac.uk/ena): Biosample: SAMEA117560877

https://www.ebi.ac.uk/biosamples

## Acknowledgments

We thank the patient and their family for participation in this study and all the clinical staff who helped with recruitment and sample collection; Dr Marina Zamudio Escalera for feedback on the manuscript; UCL IIT FACS facility for assistance with flow cytometry assays; UCL genomics for assistance with sequencing; members of all of the contributing and submitting laboratories around the globe who have openly shared large numbers of UK SARS-CoV-2 assemblies via GISAID.

## Author Contributions

DL, JB and LS conceived the project and obtained funding. JAGA, RM, KT, LF, JT, LS, DL collected and/or processed samples. JAGA performed sequencing and analysis. JAGA, RM, LS, JB, DL analysed and interpreted the data. DL, JAGA, LS prepared the manuscript. All authors reviewed and approved the manuscript.

## Materials and Methods

### Sample Processing

To monitor SARS-CoV-2 infection status over time, serial nasopharyngeal swabs were collected and tested using quantitative PCR (qPCR). A cycle threshold value ≤ 40 was considered positive for SARS-CoV-2 RNA. Samples with a positive result were sent for sequencing to determine sample lineage as previously described^12^.

### Sequencing

Libraries were hybridized to a biotinylated RNA bait set (Agilent Technologies) designed against the SARS-CoV-2 reference genome (NCBI, National Center for Biotechnology Information, GenBank MN908947.3) using the Agilent SureSelectXT Automated Target Enrichment Protocol. Captured libraries were indexed and processed using the Bravo Automated Liquid Handling Platform (Agilent). All libraries were sequenced on a MiSeq genome sequencer (Illumina) with 2×150 bp paired end reads at a target depth of 5000x coverage per genome. Positive (SARS-CoV-2 Alpha Variant RNA) and negative (nuclease-free water) controls were included in each batch of samples starting from RNA extraction through sequencing to monitor contamination.

### Sequence Analysis

Raw fastq sequencing reads were processed using fastp^52^ for adapter trimming and quality filtering using default parameters. Reads were mapped to the Wuhan-Hu-1 SARS-CoV-2 reference genome (GenBank accession MN908947.3) and the primer binding sites were trimmed from the alignment based on the ARTIC v4.1 primer scheme coordinates. Variants were called using iVAR^53^ with the default settings requiring ≥10X coverage depth. Only genomes with ≥90% coverage of the reference at 10X depth were retained for analysis. The Phylogenetic Assignment of Named Global Outbreak LINeages (PANGOLIN) SARS-CoV-2 tool^54^ was used to assess the lineage of each sample. The entire workflow from raw reads to consensus assembly and lineage typing was processed through the nf-core/viralrecon v2.6.0 pipeline.

### Sample Placement Analysis

Global sample placement analysis was performed using UShER^13^, which is a tool for inserting query sequences into a fixed reference phylogeny to determine evolutionary relationships. This was used to place the sequenced SARS-CoV-2 samples in global context using more than 15M sequences publicly available as of July 2023, which represent global SARS-CoV-2 diversity.

### Novel Variant Analysis

In the absence of samples from the early stages of infection, the programs augur and auspice within Nextstrain^55^ were then used to analyse and visualize the locations of mutations arising in the sequenced samples compared to the closest publicly available sequence. The number and positions of SNPs and indels unique to each sample were assessed to identify *de novo* mutations potentially arising during infection.

### Antibody immune escape and drug resistance

The Stanford Coronavirus Resistance Database (CoV-RDB)^23^ contains curated published data on SARS-CoV-2 neutralization susceptibility profiles for variants and individual spike mutations to monoclonal antibodies (493 mAb resistant mutations described), convalescent plasma, and vaccinee sera, as well as RdRP (RNA Dependent RNA Polymerase; 19 resistance mutations identified) and 3C-like protease (3CLpro; 113 resistance mutations identified) mutations involved in drug resistance (**Supp Table.5**). The database was queried using the list of mutations observed at day 899 of persistent infection to identify If any had been previously associated with resistance to nAb, 3CLpro or RdRP inhibitors.

### Impact of non-synonymous mutations on peptide presentation to T cells

To evaluate potential T cell immune escape mutations, experimentally validated epitopes described in Grifoni *et al*^19^ and Immune Epitope Database (IEDB^56^) which contain non-synonymous mutations arising during the persistent infection were identified and listed in **Supplementary Tables 2 and 3** for CD4 and CD8 epitopes respectively.

Where mutations fell within validated epitopes, the ancestral sequence peptide and variant peptide were compared for the likelihood that they would be naturally presented on MHC (elution rank %; rank is relative to a set of random peptides). NetMHCpan-4.1 and NetMHCIIpan-4.1 were used to predict the elution scores for peptides on MHC class I and II alleles respectively^57^, using default parameters. For validated CD8 epitopes without a known HLA-restriction, EL scores (**Supp Table.3**) can be calculated for a set of HLA supertype representatives on NetMHC pan 4.1. Where these predictions indicated a probably weak or strong binder, ‘predicted’ is given next to the HLA allele in (**Supp Table.3**). There is no equivalent representative list of class II supertypes.

For MHC class I binding, a weak binder is classed as a peptide with an EL rank <2.0% and a strong binder <0.5% rank. For MHC class II a weak binder is classed as a peptide with an EL rank <5.0% and a strong binder <1.0% rank^57^. For validated epitopes without a known HLA-restriction, EL scores can be calculated for a set of HLA supertype representatives on NetMHC pan 4.1. Where these predictions indicated a weak or strong binder, ‘predicted’ is given next to the HLA allele.

Where a mutation leads to a variant peptide with a lower predicted EL rank and HLA binding category (a strong binding peptide becoming a weak binder or unclassified, or a weak binder becoming unclassified), this is considered a negative impact on presentation and potential immune escape. Mutations can also lead to a higher binding rank which indicates they are predicted to improve peptide binding to HLA.

Variant peptides that were used in immunoassays in figure 5 are highlighted in green. E, envelope; EL, elution rank; M, membrane; NP, nucleoprotein; NSP, non-structural protein; ORF, open-reading frame; S, spike; SB, strong binder; WB, weak binder. Immunodominant was defined as being recognized in 3 or more studies in the meta-analysis of validated epitopes in^19^.

### *In silico* prediction of Immunogenicity for MHC Class I epitopes

Where mutations fell within validated epitopes, the ancestral sequence peptide and variant peptide sequences at epitopes were compared for predicted immunogenicity using a simple model available at www.iedb.org^58^. Amino acids at positions 1, 2, and C-terminus amino acid were masked and not considered for immunogenicity scores. Scores greater than 0 indicate the peptide is more likely than not to elicit an immune response, while a score less than 0 indicates the inverse.

### Samples for immune assays and isolation of PBMC

Samples from the persistently affected patient were collected under an NHS Research Ethics Committee approved protocol (04/Q0501/119) and the patient provided written, informed consent. Blood was taken in December 2023 and Nov 2024, 12 months and 22 months post-resolution of the chronic infection. T cell memory responses persisting after infection were assayed.

Peripheral blood mononuclear cells were isolated from heparinised blood samples using Pancoll (Pan Biotech) density-gradient centrifugation. Isolated PBMCs were cryopreserved in fetal calf serum (FCS) containing 10% DMSO and stored in liquid nitrogen. All T cell assays reported here were performed on cryopreserved PBMCs.

Comparator data (Figure 3) on T cell responses measured from samples taken pre-pandemic and as part of the COVIDsortium bioresource during the first wave of SARS-CoV-2 infections (March-August 2020) were previously published in^21,27^. The COVIDsortium bioresource was approved by the ethical committee of UK National Research Ethics Service (20/SC/0149) and registered at https://ClinicalTrials.gov (NCT04318314). Full study details of the bioresource (participant screening, study design, sample collection and sample processing) have previously been described^59^. Pre-pandemic samples from healthy donors were collected and cryopreserved before August 2019 under NHS Research Ethics Committee number 11/LO/0421. All participants provided written informed consent and the study conformed to the principles of the Helsinki Declaration.

### T cell assays

Full lists of the peptides contained in pools of overlapping peptides covering structural^60^ (see below for details of Spike peptides) and RTC^21,27^ proteins have previously been described (15-mer peptides overlapping by 10 amino acids, GL Biochem Shanghai, >80% purity). A list of optimal peptides corresponding to the ancestral or variant sequence (corresponding to non-synonymous mutations seen at day 899 of a persistent infection) is given in **Supp Table.6**. Optimal length peptides were purchased at >95% purity, GL Biochem Shanghai.

The following reagents were obtained through the NIH HIV Reagent Program, Division of AIDS, NIAID, NIH: Peptide Pool, Cytomegalovirus, Epstein-Barr Virus and Influenza Virus (FEC) Control, ARP-9808, and SARS-CoV-2 peptide array, Gene S NR-52402, contributed by DAIDS, NIAID.

The IFNγ ELISpot assay was performed as previously described on cryopreserved PBMCs^21^. Culture medium for human PBMCs (R10) was sterile 0.22-μm-filtered RPMI medium (Thermo Fisher Scientific) supplemented with 10% by volume heat-inactivated (1 h, 64 °C) FCS (Hyclone) and 1% by volume 100× penicillin and streptomycin solution (Gibco-BRL).

ELISpot plates (Merck-Millipore, MSIP4510) were coated with human anti-IFNγ antibodies (1-D1K, Mabtech; 10 μg ml−1) in PBS overnight at 4 °C. The plates were washed six times with sterile PBS and blocked with R10 for 2 h at 37 °C with 5% CO_2_. PBMCs were thawed and rested in R10 for 3 h at 37 °C with 5% CO_2_ before being counted to ensure that only viable cells were included. PBMCs (400,000 per well) were seeded in R10 and were stimulated for 16–20 h with SARS-CoV-2 peptide pools (2 μg/ml per peptide) at 37 °C in a humidified atmosphere with 5% CO_2_.

Internal plate controls were two DMSO wells (negative controls), concanavalin A (ConA, positive control; Sigma-Aldrich) and FEC (HLA I-restricted peptides from influenza, Epstein– Barr virus and CMV; 1 μg/ml per peptide). ELISpot plates were developed with human biotinylated IFNγ detection antibodies (7-B6-1, Mabtech; 1 μg/ml) for 3 h at room temperature, followed by incubation with goat anti-biotin alkaline phosphatase (Vector Laboratories; 1:1,000) for 2 h at room temperature, both diluted in PBS with 0.5% BSA by volume (Sigma-Aldrich), and finally with 50 μl per well of sterile filtered BCIP/NBT Phosphatase Substrate (Thermo Fisher Scientific) for 7 min room temperature. Plates were washed in double-distilled H_2_O and left to dry overnight before being read on the AID classic ELISpot plate reader (Immunospot S6 Universal M2 ELISpot Reader). Plate imaging and count settings were automated and consistent across all experiments.

The average of two DMSO wells was subtracted from all peptide-stimulated wells for a given PBMC sample and any response that was lower in magnitude than 2 s.d. of these sample-specific DMSO control wells was not considered to be a peptide-specific response (assigned value 0). Results were expressed as IFNγ SFCs per 10^6^ PBMCs after background subtraction.

### Short-term T cell peptide stimulations

Short-term peptide stimulations were performed as described previously^21^ and were used to expand T cell responses that were low magnitude *ex vivo*, so that cross-recognition of ancestral and variant peptides could be assessed.

Frozen PBMCs were thawed and washed twice with sterile PBS. When CTV dilution was measured, PBMC were resuspended in 1 ml R10 culture medium (2–10 × 10^6^ PBMCs) and 0.5 μl of 5 mM stock CTV (Thermo Fisher Scientific) was added per sample with mixing. PBMCs were stained in the dark for 10 min at 37 °C in a humidified atmosphere with 5% CO2. Ten-times volume of cold R10 was added to stop the staining reaction, and cells were incubated for 5 min on ice. Cells were washed in PBS and incubated for 5 min at 37 °C before being transferred to a new tube and were washed again in R10.

When CTV dilution was not measured alongside T cell expansion, PBMC were plated directly. PBMC were plated in 96-well plates (2–4 × 10^5^ PBMCs in 200 μl R10) and stimulated with peptide pools (2 μg/ml per peptide) for 5-10 days (5 days for peptide pools, 8-10 days for individual peptide stimulations) in R10 supplemented with 0.5 μg/ml soluble anti-CD28 antibodies (Thermo Fisher scientific) and 20 U/ml recombinant human IL-2 (Peprotech). Then, 100 μl medium was added on day 1, and 100 μl medium was removed and replaced with R10 supplemented with anti-CD28 and IL-2 as above on days 3 and 6. On the penultimate day of culture PBMCs were restimulated with peptide pools (2 μg/ml per peptide) and brefeldin A (10 μg/ml; Sigma-Aldrich). After 16–18 h restimulation, PBMCs were collected, washed in PBS and stained with fixable live/dead (Near infrared, Thermo Fisher Scientific, 1:1,000), washed in PBS, before being fixed in fix/perm buffer (TF staining buffer kit, eBioscience) for 20 min room temperature. Cells were washed in PBS and incubated in perm buffer (TF staining buffer kit, diluted 1:10 in double-distilled H2O) for 20 min at room temperature, washed in PBS and resuspended in perm buffer with saturating concentrations of anti-human antibodies for intracellular staining: anti-IL-2 PerCp-eFluor710 (Invitrogen, MQ1-17H12, 1:50), anti-TNFα FITC (BD Bioscience, MAb11, 1:100), anti-CD8α BV785 (BioLegend, RPA-T8, 1:200), anti-IFNγ BV605 (BD Biosciences, B27, 1:100), anti-IFNγ APC (BioLegend, 4S.B3, 1:50), anti-CD3 BUV805 (BD Biosciences, UCHT1, 1:200), anti-CD4 BUV395 (BD Biosciences, SK3, 1:200), and anti-CD154 (CD40L) Pe-Cy7 (BioLegend, 24-31, 1:50). Cells were washed twice in PBS and analysed using the BD LSRII flow cytometer. Cytometer voltages were consistent across batches. Fluorescence minus one (FMOs) and unstimulated samples were used to determine gates applied across samples. Data were analysed using FlowJo v.10.7 (TreeStar). Example gating shown in **Supp Figure 3**.

CTV dilution and/or staining with anti-human-IFNγ and anti-human-TNF antibodies were used to identify antigen-specific T cells. An unstimulated control well (equivalent DMSO volume to peptide wells) was included for each PBMC sample and the percentage of responding CD4+ or CD8+ cells in wells not stimulated with peptide were subtracted as background cytokine release from all peptide-stimulated wells. Polyfunctionality, defined as the number of cytokines co-produced by T cells after expansion for 10 days, was assessed using SPICE (v.6.0) and pestle (v.2.0), available at GitHub (https://niaid.github.io/spice/; **Figure 4c-d**^61^)

Boolean gating was used to identify the percentage of T cells making possible combinations of the following cytokines: IFNγ, TNF, IL-2, CD154, or expressing CD107a (marker of degranulation). Pestle was used to background-subtract the percentage of cytokine-producing cells from unstimulated wells that were run in parallel and to format data for visualization in SPICE. The proportion of T cells making a specific number of cytokines in combination is presented as pie graphs (base mean) and pie arcs represent the proportion making a given cytokine.

### Statistics and reproducibility

Data were assumed to have a non-Gaussian distribution and nonparametric tests were used throughout. For single-paired and unpaired comparisons, Wilcoxon matched-pairs signed-rank tests and Mann-Whitney U-tests were used. For multiple unpaired comparisons, Kruskal– Wallis one-way ANOVA with Dunn’s correction was used. For correlations, Spearman’s r test was used. P < 0.05 (two-tailed) was considered to be significant. Prism v.10.4.1 for Mac was used for analysis. Details are provided in the figure legends.

---

**Supplementary Figure 1:**
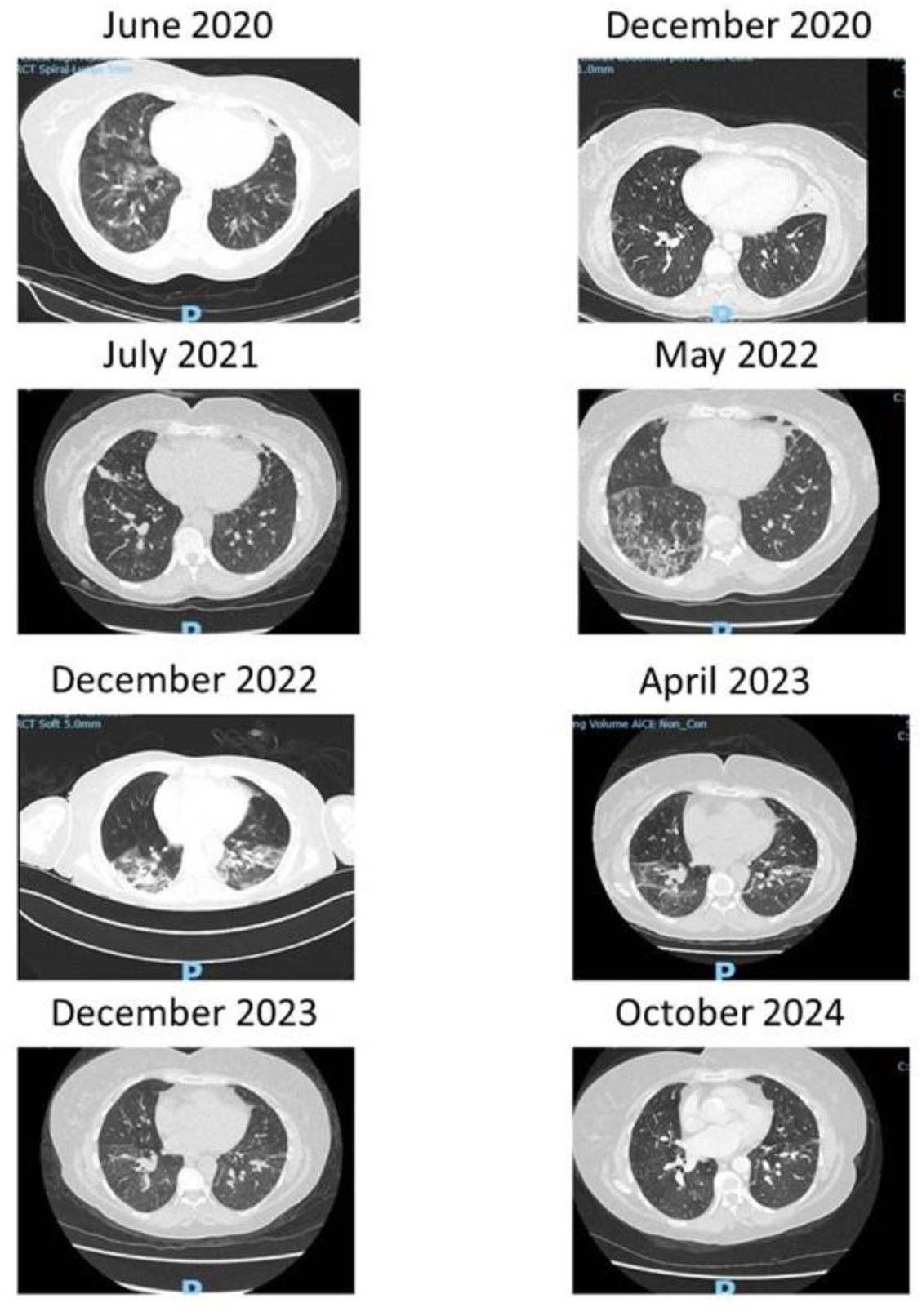
Serial chest high resolution CT imaging over time. Scans from June 2020 to December 2022 demonstrate typical ground glass changes or focal areas of consolidation, consistent with COVID-19 disease. Subsequent imaging shows resolution over time but with residual fibrosis, air trapping and large airways disease.

**Supplementary Figure 2:**
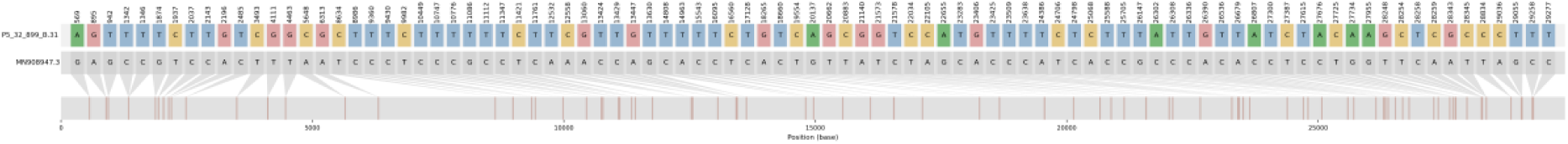
Single nucleotide polymorphisms arising compared to Wuhan hu-1 reference genome (accession number, left hand side) in the virus isolated at day 899 of persistent infection.

**Supplementary Figure 3.**
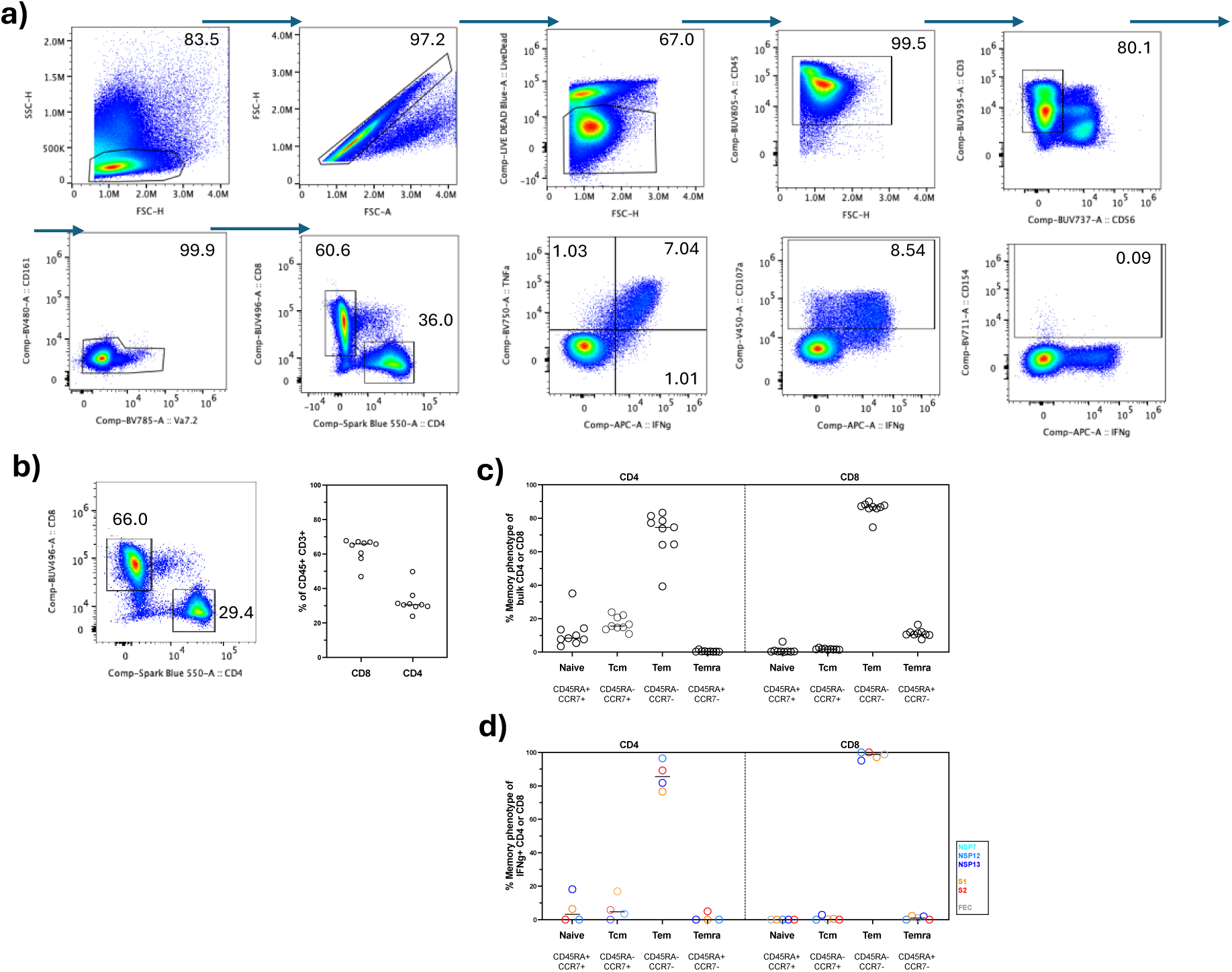
Gating strategy for intracellular cytokine staining and T cell memory phenotype: **a)** Example FACS plots showing gating on lymphocytes (SSC-H vs. FSH-H), single cells (FSC-H vs. FSC-A), live cells (Live/dead cell permeable dye negative), CD45+, CD3+CD56-(T cells), CD161^lo^, CD8+ or CD4+. Example plots of IFNγ vs. TNF, CD107a, and CD154 are shown on CD8 T cells. Stimulated with Flu, EBV, CMV epitope pool. Percentage of parent population falling in gate shown. **b)** CD4+ and CD8+ populations as a percentage of Live/CD45+/CD3+/non-MAIT T cells. **c)** Memory phenotype of bulk CD4+ and CD8+ T cells. **d)** Memory phenotype of IFNγ+ T cells after overnight stimulation with Spike region S1 and S2, NSP7,NSP12, NSP13 or a pool of immunodominant MHC class I-restricted Flu, EBV and CMV peptides. **c-d)** Bars, median. **b-c)** 9 replicate stains.

**Supplementary Figure 4:**
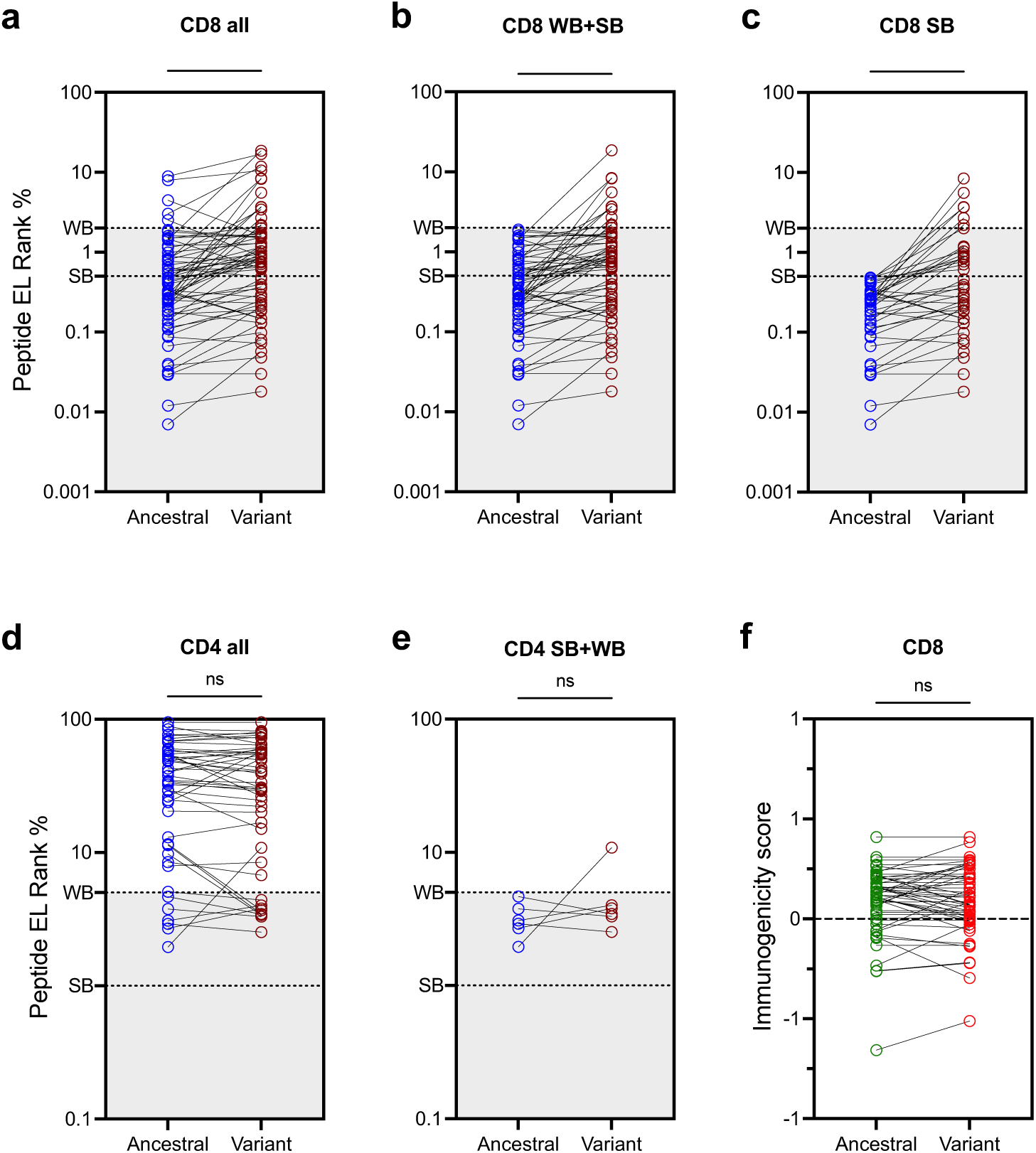
Predicted impact of mutations arising during persistent infection on peptide-MHC binding and immunogenicity. **a-e)** The percentage elution rank (calculated using NetMHCpan or NetMHCIIpan) for validated CD8+ (**a-c**; n=55) and CD4+ (**d-e**; n=38) epitopes affected by non-synonymous mutations arising during this persistent infection, comparing ancestral sequence to variant sequence. **a,d**) all epitopes, (**b,e**) only those with ancestral sequence predicted as weak or strong binders (WB+SB), **c**) only strong binders. All validated HLA allele-peptide combinations are included. f) *In silico* predicted Immunogenicity scores of all CD8 epitopes affected by mutations arising during this persistent infection, comparing ancestral and variant sequence. Statistical analysis was performed using Wilcoxon paired non-parametric t-test. Epitope lists are given in **Supplementary Tables 2&3**.

**Supplementary Table 1: Sequencing quality control metrics for SARS-CoV-2 samples.** This table summarizes sequencing quality metrics for all SARS-CoV-2 samples obtained from the patient. Quality control status was determined as “Pass” for samples with ≥90% genome coverage at 10X depth or greater, and “Fail” for samples below this threshold. Only samples with “Pass” status were used for downstream phylogenetic and mutation analyses.

**Supplementary Table 2: List of validated CD4+ T cell epitopes in which mutations arising during persistent infection fall and predicted impact on peptide HLA binding.** Variant peptides that were used in immunoassays in Fig. 5 are indicated in ‘Tested in immune assays column’. Partial means likely overlap of core peptide region but not exact peptide match with that used in immune assays. E, envelope; EL, elution rank; M, membrane; NP, nucleoprotein; NSP, non-structural protein; ORF, open-reading frame; S, spike; SB, strong binder; WB, weak binder. ^ Immunodominant defined as being recognized in 3 or more studies in the meta-analysis of validated epitopes in (Grifoni *et al*^19^).

**Supplementary Table 3: List of validated CD8 epitopes in which mutations arising during persistent infection fall and predicted impact on peptide HLA binding.** Where a mutation leads to a variant peptide that is predicted to impact presentation (a change of classification of WB, SB, unclassified) the EL rank scores are coloured green then red when the mutation leads to a loss of predicted presentation and red then green when the mutations leads to a higher predicted presentation category. Variant peptides that were used in immunoassays in Fig. 5 are indicated in ‘tested in immune assays’ column. E, envelope; EL, elution rank; M, membrane; NP, nucleoprotein; NSP, non-structural protein; ORF, open-reading frame; S, spike; SB, strong binder; WB, weak binder. ^ Immunodominant defined as being recognized in 3 or more studies in the meta-analysis of validated epitopes in (Grifoni *et al*^19^). For validated epitopes without a known HLA-restriction, EL scores can be calculated for a set of HLA supertype representatives on NetMHC pan 4.1. Where these predictions indicated a probably weak or strong binder, ‘predicted’ is given next to the HLA allele. * indicates no predicted HLA binders in supertype representatives.

**Supplementary Table 4: Predicted impact of mutations on peptide immunogenicity:** Immunogenicity scores were predicted using an immunogenicity model, masking anchor residues to assess peptide changes independent of potential impacts on MHC binding (Calis *et al*^58^). ▵ score denotes the difference between ancestral and variant peptide immunogenicity scores. Green denotes increase in predicted immunogenicity and red decrease for variant peptide relative to original ancestral sequence.

**Supplementary Table 5: Lists of mutations associated with resistance to mAb, protease inhibitors or RNA-dependent RNA polymerase inhibitors taken from Stanford coronavirus database (CoV-RDB).** None of the mutations listed here are observed in the virus isolated at day 899 of persistent infection.

**Supplementary Table 6: Peptides used in immune assays.** The amino acids that are substituted in the variant viral sequence isolated at day 899 of infection are highlighted in bold. Peptides used at >95% purity. See **Supplementary Tables 2 & 3** for full details on the epitopes considered, such as HLA restriction and immunodominance.

